# Quality of diabetes care at a Ghanaian Teaching Hospital (2022–2023): Retrospective chart review benchmarking against American Diabetes Association (ADA) Standards

**DOI:** 10.64898/2026.08.04.26359656

**Authors:** Daniel Kwabena Amoako-Adjei, Paul Nsiah, Evans Ansu-Yeboah

## Abstract

**Background:** Diabetes mellitus is a global major health concern, with increasing prevalence in Ghana, causing significant mortality and morbidity. Effective management of this chronic condition depends on resilient healthcare systems that adhere to evidence-based guidelines. This study aimed to evaluate the quality-of-care at the diabetic clinic of the Cape Coast Teaching Hospital (CCTH) using the American Diabetes Association (ADA) guidelines as the benchmarks to show the performance of the hospital clearly, reveal gaps in the management algorithm that need to be addressed, and to contribute to the body of knowledge in the health sector of our country.

**Methods:** Retrospective chart review was done from January 2022 to March 2023. 261 patient records were used out of a total study population of 780. Primary process indicators (medical history, physical examination, laboratory check-ups, referrals) and outcome indicators (HbA1c<7%, BP<140/90 mmHg, lipid profile, liver function test, kidney function based on eGFR categories). In analyzing the data, the Statistical Package for Social Sciences (SPSS version 21) software and STATA vs.14 were used to generate frequencies, means, and percentages presented in tables, charts, and graphs to provide answers to the research questions.

**Results:** The overall documented adherence rates were: medical history (17%), physical examination (58.4%), laboratory evaluation (24%), and referrals (5%). These were coupled with low rates of achievement of treatment targets for various clinical outcomes, specifically glycated hemoglobin (HbA1c), blood pressure, lipid profile and estimated glomerular filtration rate (eGFR), with corresponding percentages of 47.2%, 42%, 47.1%, and 48.8%, respectively.

**Conclusion:** The study, therefore, revealed gaps in compliance and documentation, which should be addressed to improve clinical care and patient outcomes.

## Introduction

Diabetes mellitus is a major global health concern with increasing prevalence, causing significant mortality and morbidity [1]. According to the International Diabetes Federation (IDF) Diabetes Atlas, Tenth edition 2021, about 537 million adults (20-79 years) are living with diabetes, and this number is projected to rise to 643 million by 2030 and 783 million by 2045. The disease burden is disproportionately high amongst adults in low- and middle-income countries (about 75% of cases), with almost half remaining undiagnosed [2]. In Sub-Saharan Africa (SSA), the awareness, treatment, and control rates are low, contributing to severe complications of the disease and premature mortality, as 75% of diabetes related deaths in SSA occur before age 60 [3]. Local statistics within Ghana, a sub-Saharan African country, have revealed an increase in the disease prevalence due to demographic changes, transition in culture, and other factors (4 ,5). These depressing statistics concerning DM in SSA reflect a substandard architecture for DM care in SSA, which demands urgent strengthening and evaluation [3].

One major challenge amongst patients with diabetes mellitus in Africa is that inhabitants who have the condition may be ignorant of it until severe complications develop [6]. A healthcare system that undertakes comprehensive medical evaluation and assessment of comorbidities is the solution to these challenges of late diagnosis and late presentation with complications. The World Health Organization (WHO) and the American Diabetes Association (ADA) have clearly emphasized the relevance of evidence-based guidelines in reducing or eliminating microvascular or macrovascular complications of DM which contribute to the increased mortality and morbidity associated with the disease (1,7).

The increasing prevalence of this chronic disease in Ghana calls for review and periodic evaluation of our guidelines for diabetes care in its adherence and conformity to international standards of care to help promote diabetes care (5). The current guideline that was being used by the diabetic clinic of the Cape Coast Teaching Hospital (CCTH), as at the time of the research, was prepared from two resources which are the Ghana Standard treatment guidelines (STGs) and the ADA guideline as Ghana had no established national comprehensive guideline for diabetes care only. However, no evaluation has been done to this date to assess the clinicians’ adherence to these guidelines and also patient outcomes. Comparative analysis of national guidelines for management of chronic diseases across low, middle and high-income countries revealed there is no strength and evidence in the recommendations the Ghana STGs provide on management of type 2 DM relative to guidelines used in high income countries such as those of the American Diabetes Association [8]. The American Diabetes Association (ADA) guidelines are evidence-based guidelines based on large randomized clinical trials which were done using participants from high-income countries (HICs) but there is paucity of such data and research in our Sub-Saharan region and so there is hardly any comprehensive management guidelines that fit our context [9]. Moreover, adherence to several aspects of the ADA management recommendations has been shown to reduce many of the complications of diabetes [10].

There is paucity of data available on the evaluation of diabetes care in Ghana as compared to studies on the prevalence and risk factors, and this calls for the need for such research to be done to make more data available to inform policy making and planning of healthcare to improve management outcomes.

The third sustainable development goal of the United Nations is ensuring healthy lives and promoting wellbeing at all ages, and one of the targets under this goal is to reduce by one-third premature deaths that arise from non-communicable diseases (NCDs) and this is a global call to strengthen health management systems, especially in SSA [11]

This study, therefore, sought to evaluate the documented adherence of the diabetic clinic at CCTH to ADA guidelines and its impact on patient outcomes.

## Materials and methods

### Study design

A retrospective cross-sectional study was done using a checklist constructed based on the American Diabetes standards (ADA) of care, 2021.

### Study setting

This study was conducted in the Cape Coast Teaching Hospital, a tertiary-level hospital that serves as a referral site for all the district hospitals in the Central Region, as well as the Winneba Trauma Centre. All patient records are computerized in the hospital. The diabetic clinic is run by the internal medicine department. The clinic is staffed by nurses and physicians. Attending physicians at the clinic are usually medical officers (general practitioners) and sometimes internal medicine specialists.

### Study population

Target population were diabetic patients who received care at CCTH Diabetes clinic between January 2021 and March 2023 and the number of such patients according to the medical records unit of the hospital was 780 and out of this, a sample of 261 patients was selected. The research sought to evaluate what is currently being done in the management of diabetic patients and that is why data from more recent years were chosen with inclusion and exclusion criteria clearly stated below.

### Eligibility criteria

All patient records used for the study had to meet all these 3 conditions which included; outpatients who have received care at CCTH, male or female clients above 18 years who had been diagnosed with diabetes for at least 1 year with active follow up at CCTH. A single exclusion criterion amongst the criteria stated below was sufficient to exclude a patient record from the study. The criteria included patients visiting the diabetic clinic for the first time who had records of only their first visit, patients who had only inpatient records with no outpatient record of a diabetic clinic encounter, and all pregnant women.

### Study hypothesis

This study hypothesized that there is a high documented adherence to ADA standards with no significant difference or gaps.

### Sampling procedure

A sample size of 258 was calculated using the Cochran’s formula which is considered appropriate in situations with large populations as seen in this research. Patients who had been diagnosed with diabetes for at least 1 year and who received care at the CCTH Diabetes clinic between January 2021 and March 2023 and who meet the selection criteria were randomly selected from the electronic database of CCTH. The list of patients, 780 in total, was transformed from the manually written database at the diabetic clinic into an Excel format.

Data from the Excel format was then imported into STATA vs 14 where simple random sampling was done to get the calculated sample size.

### Data collection instrument

A checklist, constructed based on the American Diabetes standards (ADA) for diabetes care 2021, was used as a guide to obtain data. The checklist was based on four main domains; medical history, physical examination, laboratory check-ups, and referrals to other clinics. Medical records were examined for evidence of documentation of the various clinical parameters or variables.

The variables of focus for medical history were hypoglycaemia episodes, visits to specialists, eating patterns and weight history, physical activity and sleep behaviours, current medication regimen, medication-taking behaviour, medication intolerance or side effects, complementary and alternative medicine use.

Those for physical examination were height, weight and body mass index (BMI), blood pressure determination and skin examination.

Those for laboratory evaluation were HbA1c, lipid profile, liver function test, spot urinary albumin-to-creatinine ratio, serum creatinine and estimated glomerular filtration (eGFR) rate, thyroid stimulating hormone, vitamin B12 if on metformin, serum potassium levels in patients on ACE inhibitors, ARBs or diuretics.

Referrals were also considered and those used in the study were eye care professional for annual dilated eye exam, registered dietitian nutritionist for medical nutrition therapy, diabetes self-management education and support, dentist for comprehensive dental and periodontal examination.

### Data collection procedures

A list of all diabetic patients who received care at CCTH Diabetes clinic between January 2021 and March 2023 alongside their electronic codes for access to their electronic records was generated electronically from the manually written database into an Excel file which was then imported to STATA for a simple random sampling to be done through which we arrived at the sample. Patients in the selected sample had their records viewed from the hospital’s electronic database using the checklist generated between April 10, 2023, and May 31, 2023. Primarily, only follow-up visits, after the first initial visit of every client to the facility, were assessed for data collection. The medical history, physical examination and treatment parameters were all assessed using the first and last clinic visits in 2022, which were all follow-up visits, as the index visits or visits of focus. However, for the laboratory check-up and referral parameters, all visits from the beginning of 2022 to March 2023 were used in its evaluation and data collection. The following clinical outcome measurements were also retrieved; blood pressure, fasting plasma glucose, random plasma glucose, renal function test results, lipid profile, glycated haemoglobin and liver function test results. These results were the most recent ones available from January 2022 up to March 2023. Information retrieved from patient records was captured and compiled in an excel file which was then imported to SPSS for data analysis. No patient contact or testing was performed during this study.

Any missing data in the patient’s records pertaining to the variables of interest in the study was considered as ‘not done’ as a more conservative approach. Documentation was used as the only objective proof that a particular process or activity was carried out, and for any process duly carried out, it was marked with a YES on the data collection Excel sheet, but for those not done, it was represented with a NO on the data collection Excel sheet.

For the laboratory parameters, tests ordered regardless of the availability of results were marked as Yes and deemed compliant with the ADA standards. For specific labs which did not apply to all patients, NA (not applicable), was used to represent such cases.

Compliance rates were therefore a reflection of the percentage of persons who were taken through the various performance indicators as indicated in the checklist. Two follow-up visits were used to assess the history and physical examination quality indicators, while all visits from the beginning of 2022 to March 2023 were used to assess the laboratory parameters, as well as the referrals within the context of what is required per the ADA guidelines for every follow-up visit, as well as annual visits.

All data were anonymized and individual identifiers were removed prior to analysis to protect participant confidentiality.

### Data processing and analysis

Medical records of 261 patients who met the inclusion criteria were included in the analysis. Data abstracted from the electronic database was entered into Statistical Package for Social Sciences (SPSS version 21) and crosschecked to ascertain data errors. Discrepancies and consistency checks of all the data entered were done. For variables which had missing data, such data was excluded from analysis.

Analysis of the data obtained was done using the Statistical Package for Social Sciences (SPSS) version 21.0 and Microsoft Excel. Data was de-identified and presented in an aggregated format. Descriptive statistics were used to briefly describe the sample. Frequencies, percentages, mean and ranges have been generated and appropriately represented by tables, graphs and charts in the write-up to provide answers to research questions thereby meeting the objectives of the study. A simple logistic regression analysis was also used to assess if blood pressure measurements done for every follow-up visit and annual HbA1c measurements were statistically significant predictors of blood pressure control outcomes and glycaemic control outcomes, respectively.

### Ethical consideration

Ethical clearance was given by the CCTH Institutional Review Board (IRB). The IRB approved a waiver of informed consent due to retrospective review of de-identified charts (Protocol number: CCTH/ERC/EC/2022/051).

## Results

### Sample characteristics

The mean age was 63 years (**±** 10.16), and the age category with the highest frequency was 61 to 80 years. The females made up a greater proportion relative to the males as shown in Table 1 below.

**Table 1.**
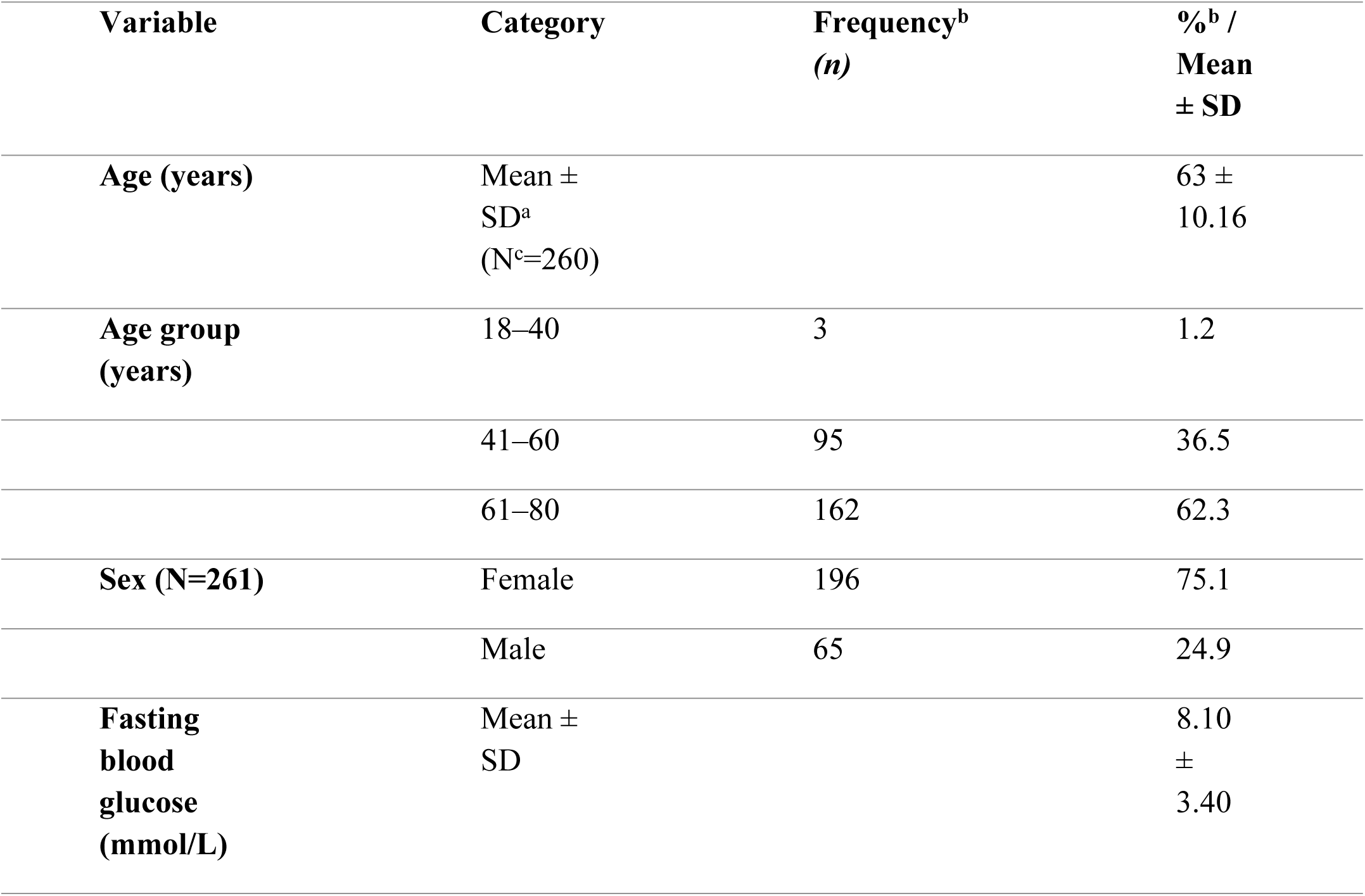
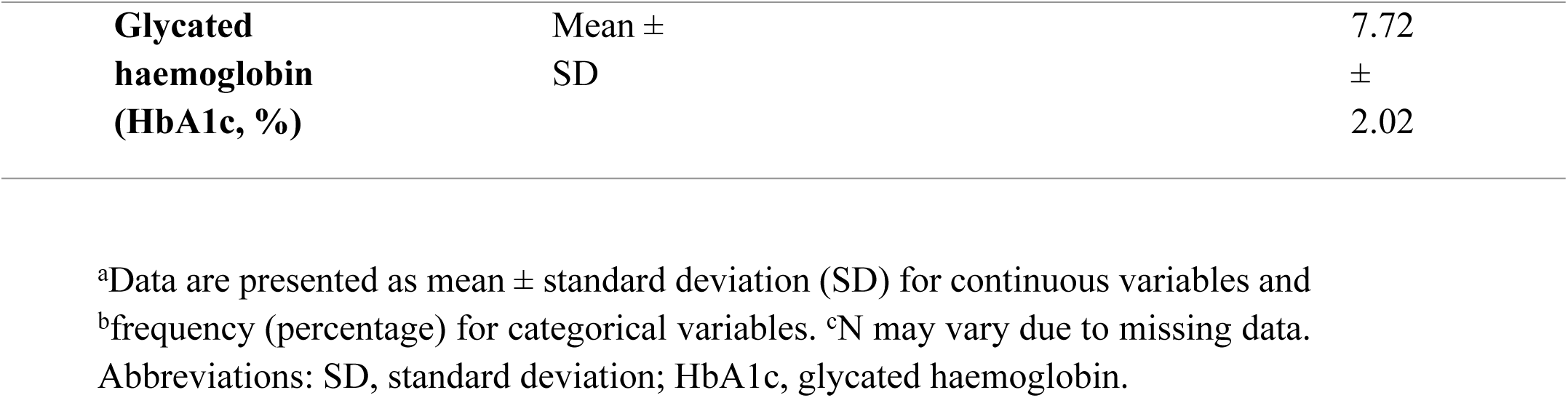
Sociodemographic and clinical characteristics of the study population.

| Variable | Category | Frequency<sup>b</sup><br>(*n*) | %<sup>b</sup> /<br>Mean<br>± SD |
| --- | --- | --- | --- |
| Age (years) | Mean ±<br>SD<sup>a</sup><br>(N<sup>c</sup>=260) | | 63 ±<br>10.16 |
| Age group<br>(years) | 18–40 | 3 | 1.2 |
|  | 41–60 | 95 | 36.5 |
|  | 61–80 | 162 | 62.3 |
| Sex (N=261) | Female | 196 | 75.1 |
|  | Male | 65 | 24.9 |
| Fasting<br>blood<br>glucose<br>(mmol/L) | Mean ±<br>SD | | 8.10<br>±<br>3.40 |

|  |  |  |
| --- | --- | --- |
| **Glycated**<br>**haemoglobin**<br>**(HbA1c, %)** | Mean ±<br>SD | 7.72<br>±<br>2.02 |

<sup>a</sup>Data are presented as mean ± standard deviation (SD) for continuous variables and <sup>b</sup>frequency (percentage) for categorical variables. <sup>c</sup>N may vary due to missing data. Abbreviations: SD, standard deviation; HbA1c, glycated haemoglobin.

### Assessment of care based on history taking, physical examination, referrals and laboratory evaluation

Adherence to ADA-recommended medical history documentation was remarkably low, with an overall documented compliance rate of 17%. Upon review of all the sampled records, hypoglycaemia assessment was remarkably low per documentation, and enquiry about visits to specialists was barely done in history taking by the clinicians as shown in Table 2 below. The study revealed that physicians hardly considered behavioural patterns in their interaction with patients during history taking, and even if they did, it was not documented to provide objective evidence to back it up.

**Table 2.**
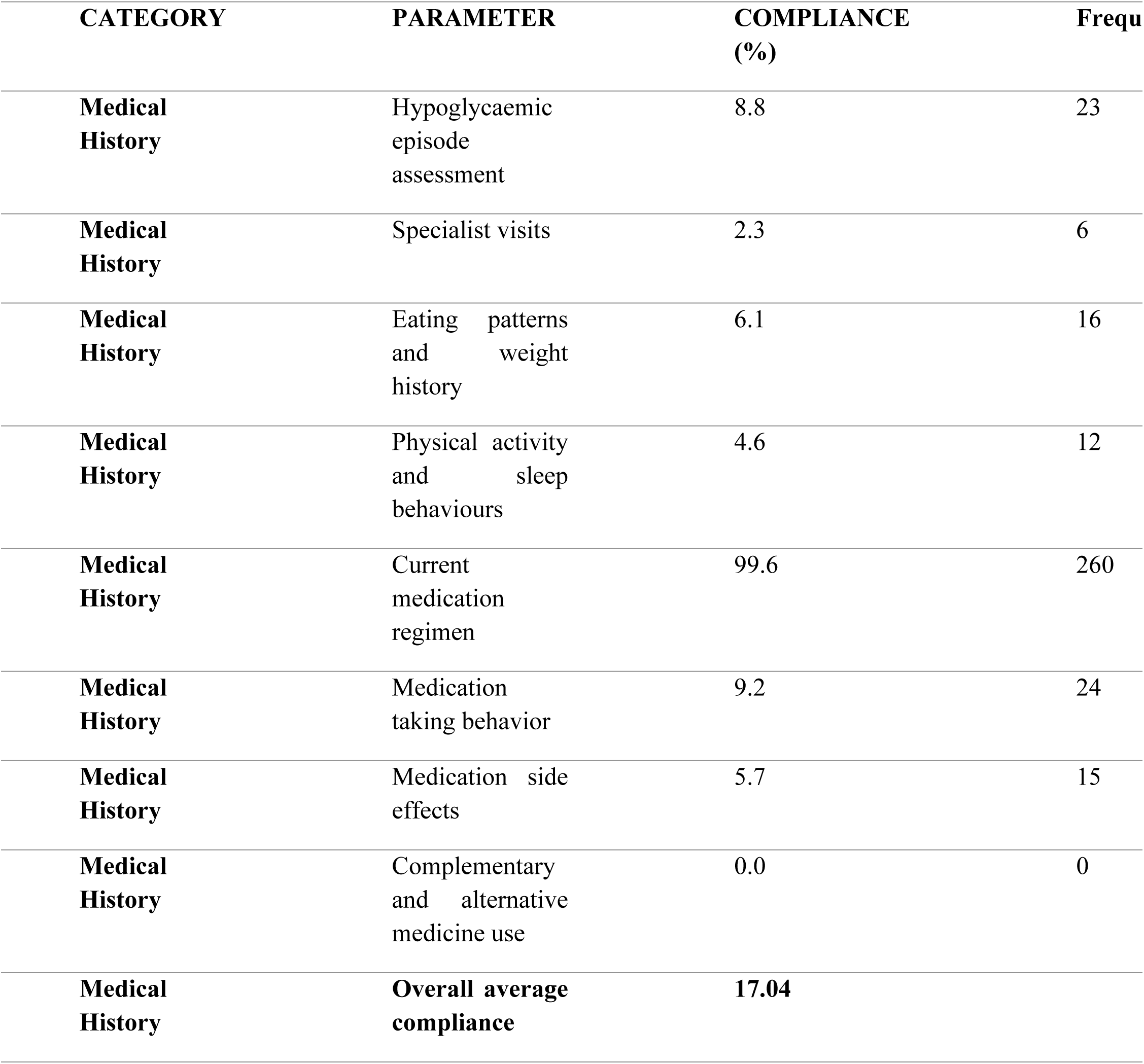
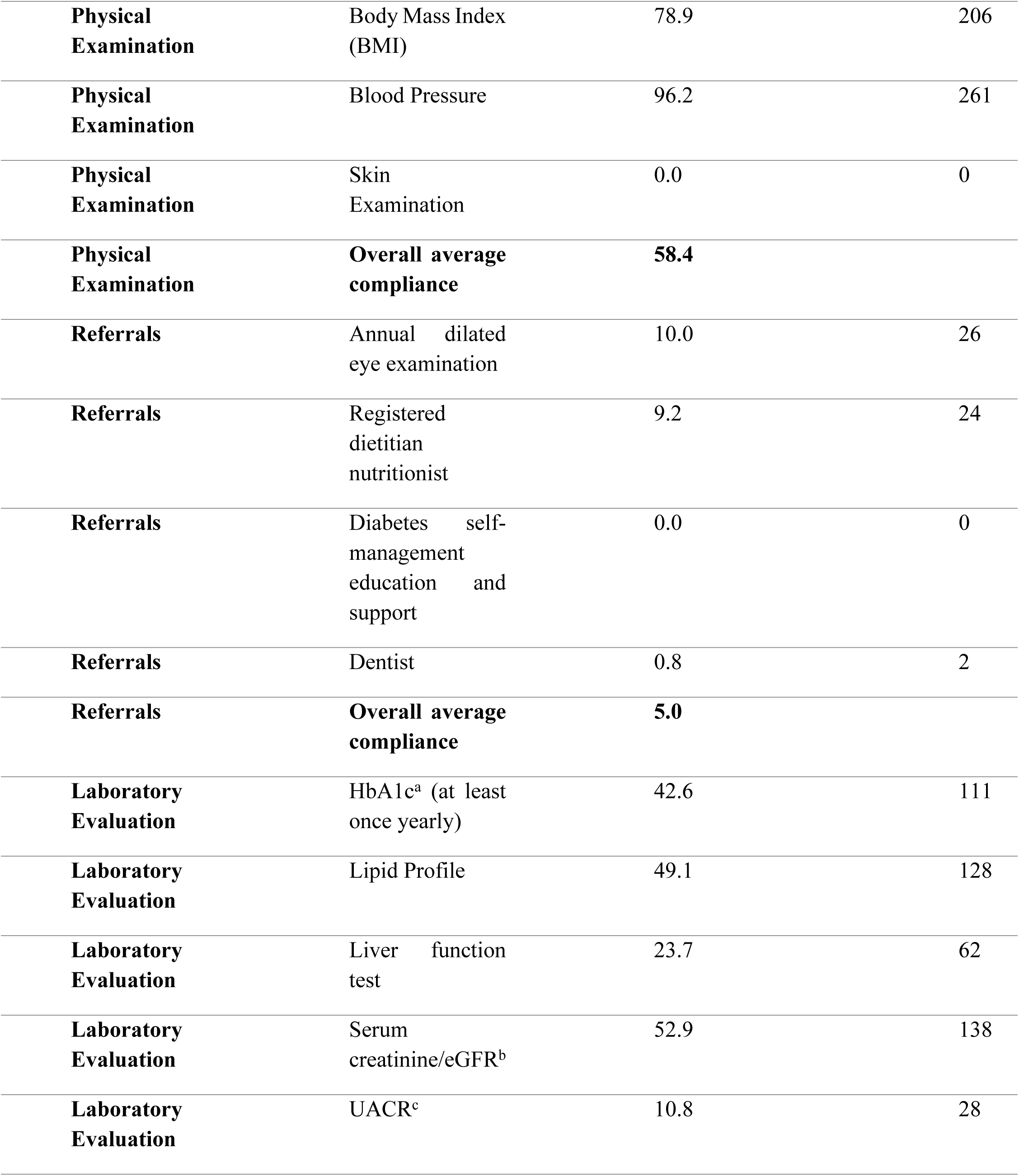
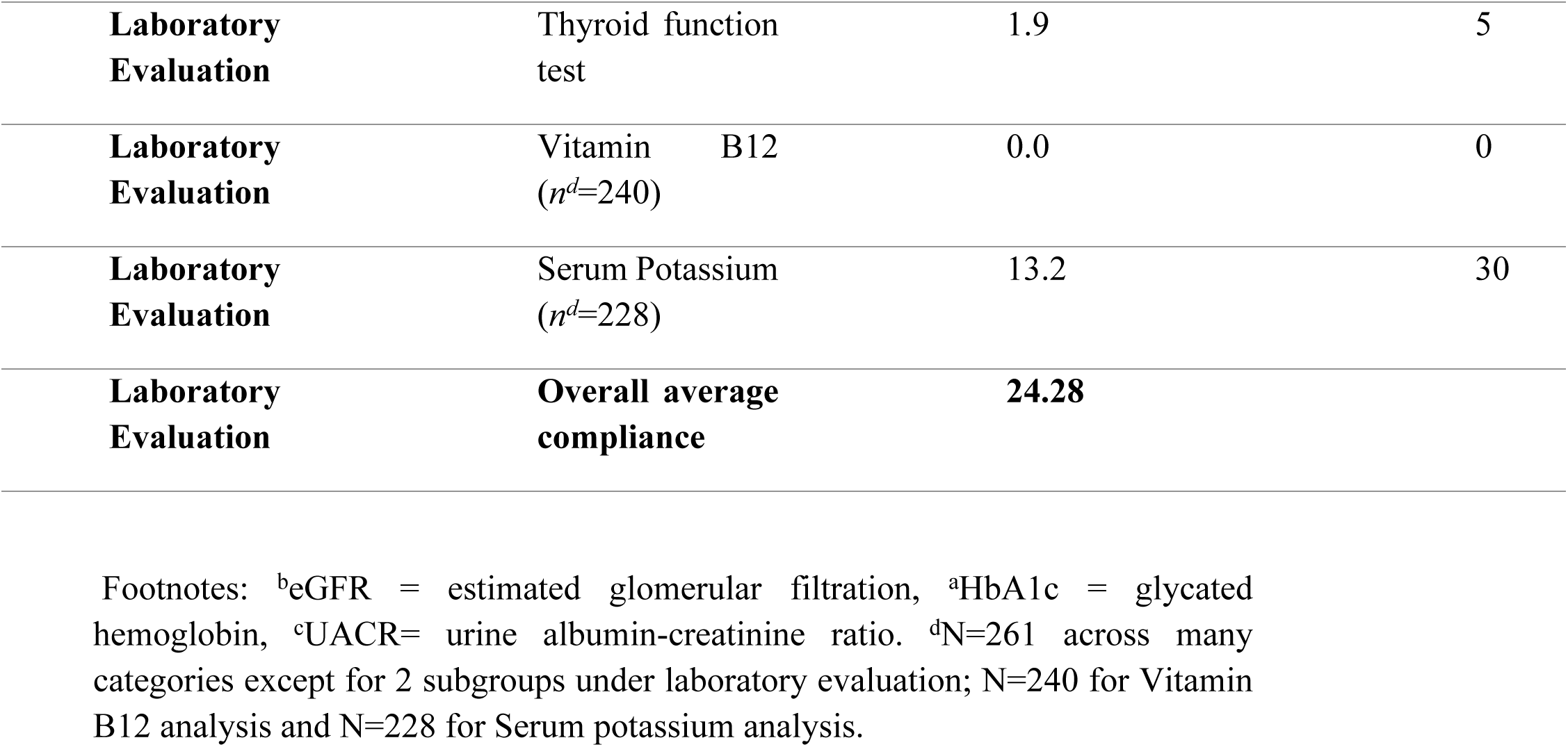
Adherence Rates Across Several HealthCare Parameters (*N^d^*= 261)

Physical examination adherence was relatively higher, with an overall compliance rate of 58.4% as depicted in Table 2. The performance of healthcare givers at the diabetic clinic in carrying out blood pressure measurement as well as height and weight measurement was good, but very poor for skin examination. There was no record of skin examination being done for any of the patients in the sample for the study.

Referral practices were the weakest areas of performance, with an overall compliance rate of 5%. As shown in Table 2, referral to the ophthalmology clinic had the highest frequency followed by referral to the dietician.

According to the ADA guidelines, some laboratory investigations are supposed to be done on annual visits, which suggests that they should be ordered for and done by every diabetic at least once a year, and these were assessed as well. Laboratory evaluations showed an overall compliance rate of 24% as shown in Table 2 below.

The performance in carrying out these investigations was not the best, as the percentage of records in which these tests were not ordered at all per documentation was above 50% for all the investigations, with the exception of serum creatinine, with a percentage of 47.1% which is also significant.

According to ADA standards, some of these tests are associated with certain conditions; for instance, vitamin B12 is ordered only for patients on metformin, and serum potassium for patients on angiotensin converting enzyme inhibitors, angiotensin receptor blockers, or diuretics, and so for patients who were not on these medications, these particular tests were not applicable to them. For urinary albumin to creatinine ratio, thyroid function test, vitamin B12, and serum potassium, the percentage of patients in which these tests were ordered was all low.

### Clinical outcomes

According to various studies, including the United Kingdom Prospective Diabetes Mellitus study (UKPDS), achieving HbA1c targets less than 7% has been shown to reduce the microvascular complications of diabetes mellitus [12], and that is why, per the ADA standards, an HbA1c target of less than 7% for many non-pregnant women without substantial hypoglycaemia is considered appropriate [12]. Results less than or equal to the target goal of 7% were considered well-controlled, while those beyond were classified as poorly controlled. It is evident from Table 3 that poorly controlled HbA1c made up the majority (52.8%).

**Table 3.**
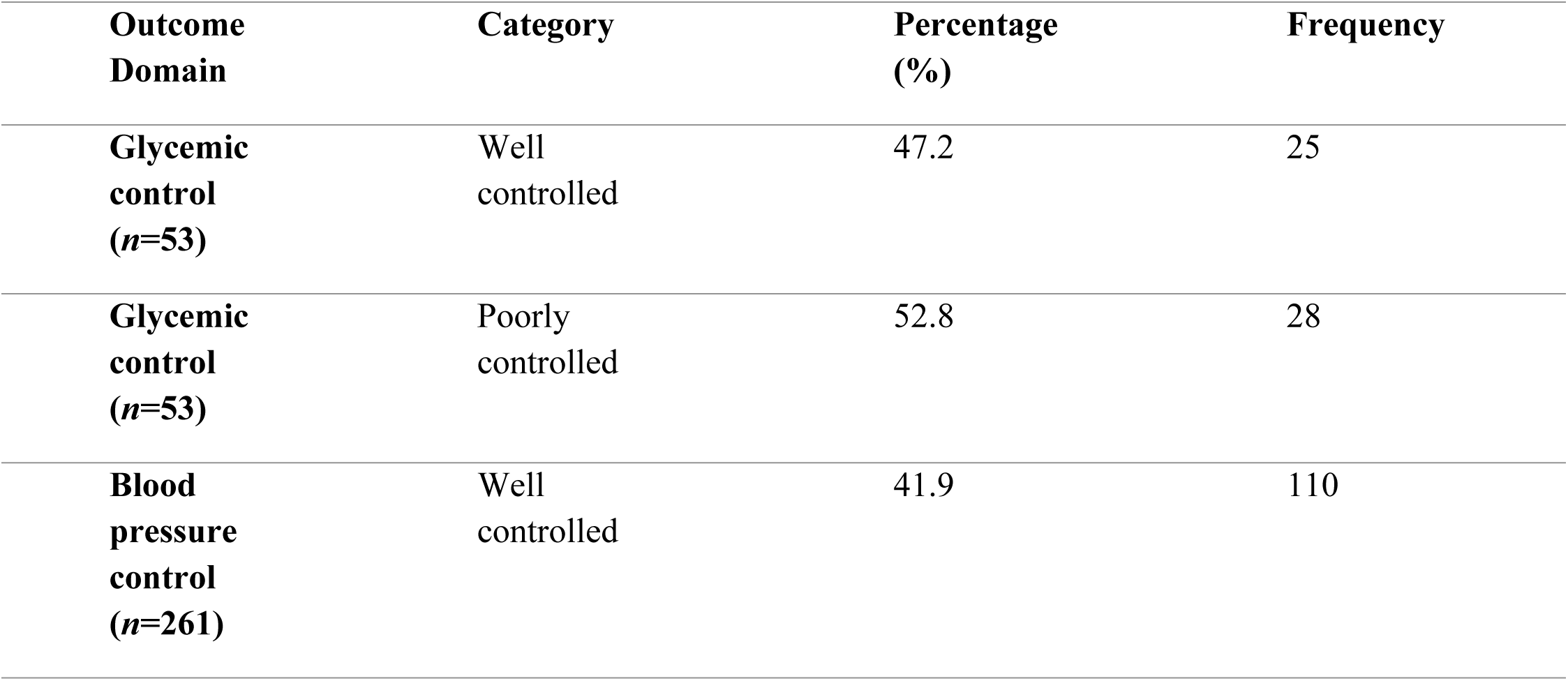
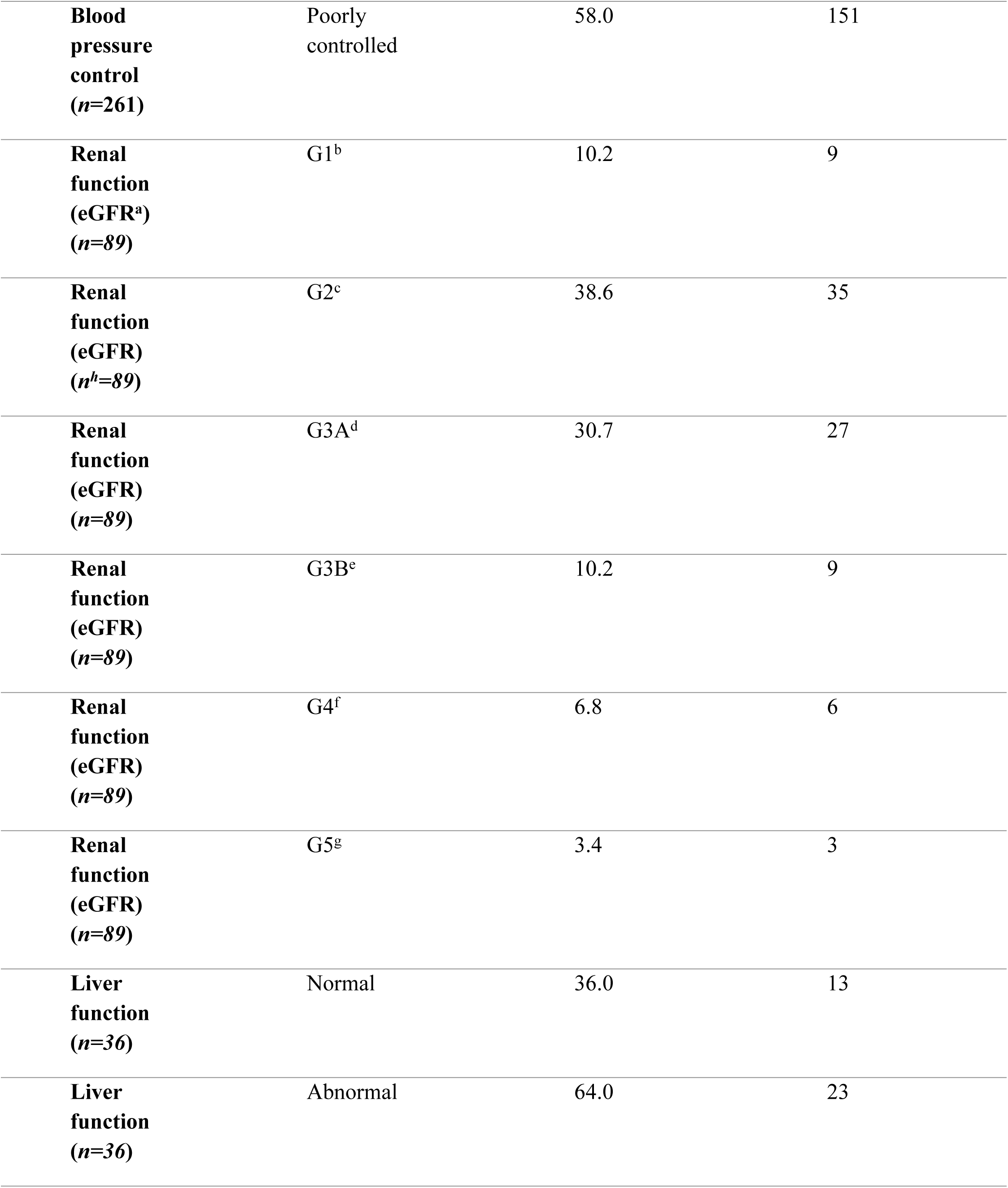
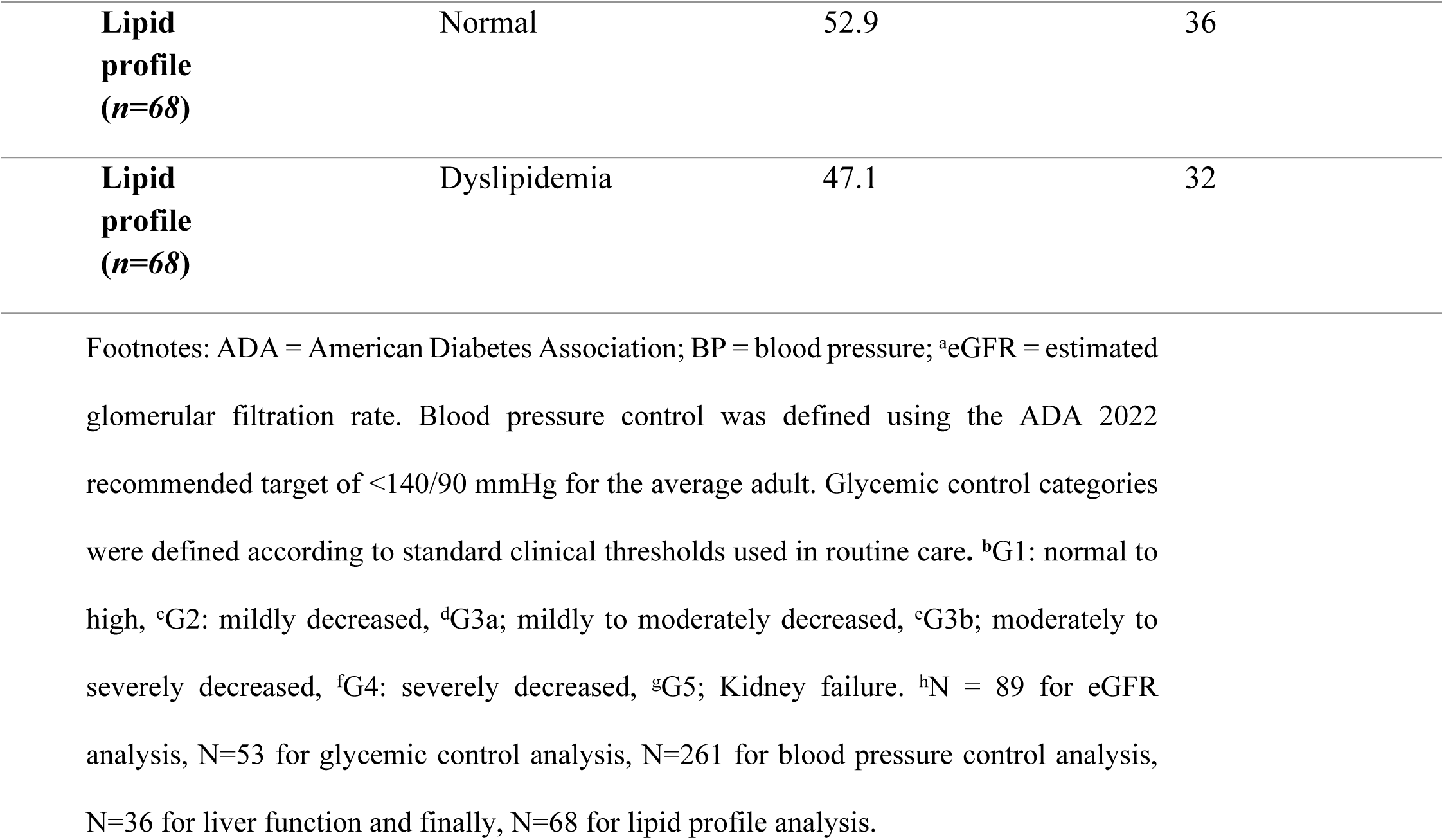
Clinical outcomes among adult patients with diabetes receiving routine care.

Randomized clinical trials have shown clearly that the treatment of hypertension to a blood pressure target less than 140/90 mmHg markedly reduces adverse cardiovascular events as well as microvascular complications of diabetes [12]. The blood pressure level of 140/90 mmHg was therefore used as the baseline for determining whether blood pressure was well controlled or not, and the majority (58%) of the most recent blood pressure recordings up to March 2023 from the records viewed were beyond the target value, thus poorly controlled.

Amongst the 261 records reviewed, no recent renal function test result from the beginning of 2022 till March 2023 was available for 172 patients. For the remaining patients with uploaded results on the electronic record system, the estimated glomerular filtration rates were categorized according to the Kidney Disease Improving Global Outcomes (KDIGO) system of classification as shown in Table 3 below. G2 (mildly decreased eGFR) had the highest frequency amongst the available results and G5 had the lowest. According to the 2022 KDIGO guidelines, chronic kidney disease (CKD) is defined as a persistent rise in urine albumin excretion, a decreased eGFR (<60 ml/min per 1.73 m^2^) that persists for more than 3 months [13]. Therefore, for this study, eGFR above 60(G1 and G2) was considered normal, as kidney function is not lost significantly, and the percentage of patients achieving this range was 48.8%.

About 73.5% of the records viewed had no uploaded lipid profile results on the information system between January 2022 and March 2023; however, out of the recent uploaded results, which were readily available, approximately 53% had a deranged lipid profile as depicted in Table 3. Dyslipidaemia is simply the imbalance of lipids such as triglycerides, LDL-cholesterol, and HDL-cholesterol, amongst others. Low HDL-cholesterol was the most common abnormal specific marker recorded.

There were also no available liver function results between January 2022 and March 2023 for about 86% of the records viewed, but out of the uploaded results, which were readily available, about 64% of patients had abnormal values for at least one of the various specific markers, and the most common derangement observed was a high GGT. This is evident in Table 3 as shown below.

### Inferential statistics

A simple logistic regression analysis was done to find out if adherence to some specific ADA process standards is a substantive predictor variable of patient outcomes in relation to those particular process standards. Table 4 reveals the outcome of these analyses.

**Table 4.**
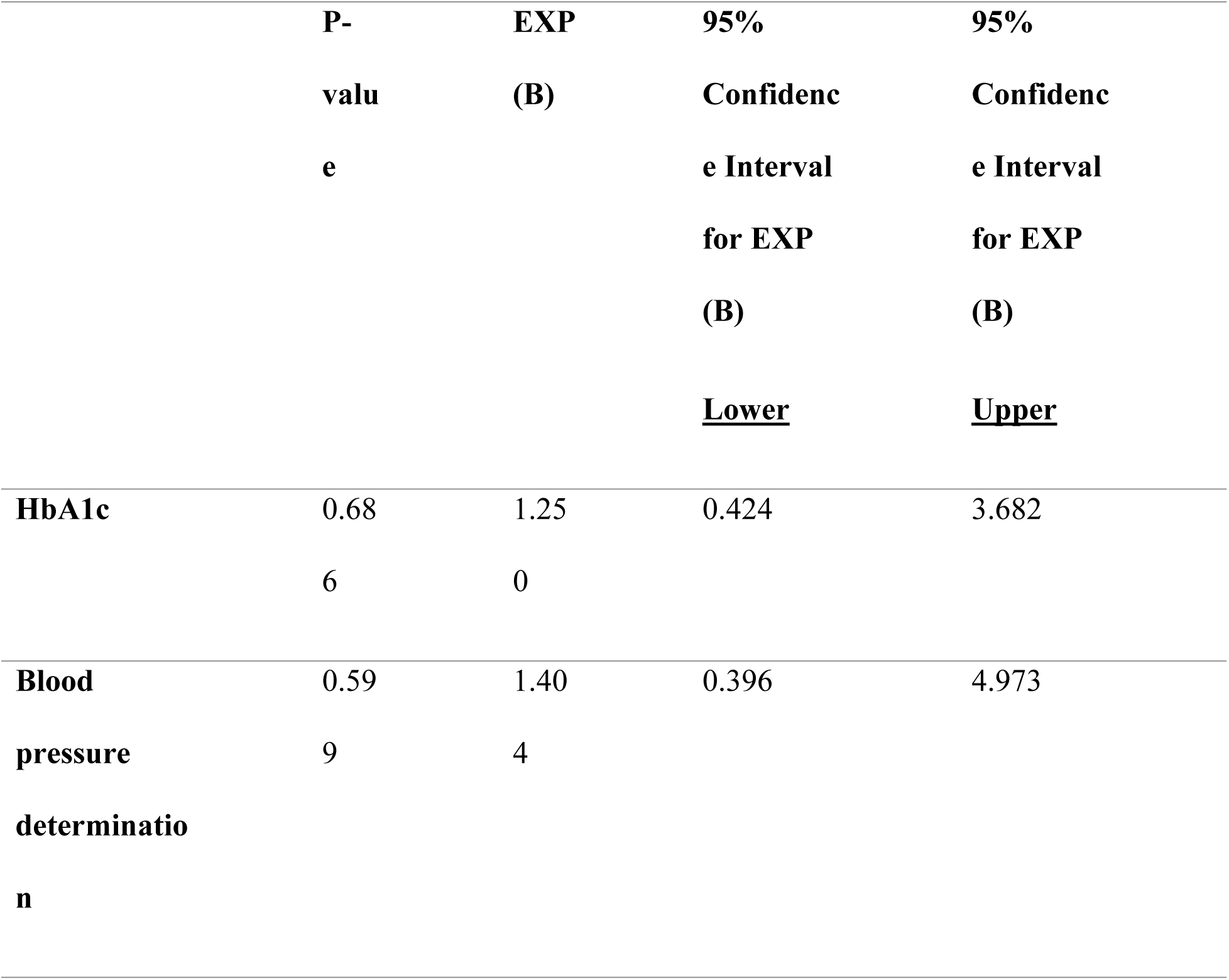
Blood pressure and HbA1c determination as predictor variables for blood pressure and glycemic control outcomes, respectively.

The odds of a well-controlled blood pressure in those who did not have their blood pressure measurement taken is about 1.4 times the odds for those whose blood pressure measurement were taken and recorded. This signifies that the probability of having a well-controlled blood pressure for patients, whose blood pressures were not determined during follow up patient visits, is higher than for patients whose blood pressure measurements were taken and recorded during patient follow up visits. However, a p value of 0.599 which is greater than the significance level of 0.05 makes the findings statistically insignificant.

According to Table 4, the odds of having a poor glycaemic control amongst those patients who did not have their HbA1c checked were 1.25 times higher than in those who had their HbA1c checked in the year 2022, but with no statistical significance, p value > 0.05.

The section on clinical outcomes reveals that diabetic patients are not doing well under the system of care, as the highest percentages were always recorded for abnormal or poorly controlled results. It should, however, be noted that due to poor documentation and upload of laboratory investigations, a significant amount of results were not available to be included in the analysis, which may mask the real picture as compared to the results obtained. The simple logistic regression analysis done also revealed that blood pressure measurements during follow up clinic visits and annual HbA1c measurements were not statistically significant predictors of how well blood pressure control and glycaemic control is achieved in diabetic patients.

Table 5 below shows that having eating patterns and weight history, physical activity and sleep behaviors, medication-taking behaviors, and weight, height, and BMI recorded by the attending physician were not significant predictors of HbA1c among patients suffering from diabetes at the diabetic clinic at Cape Coast Teaching Hospital.

**Table 5.**
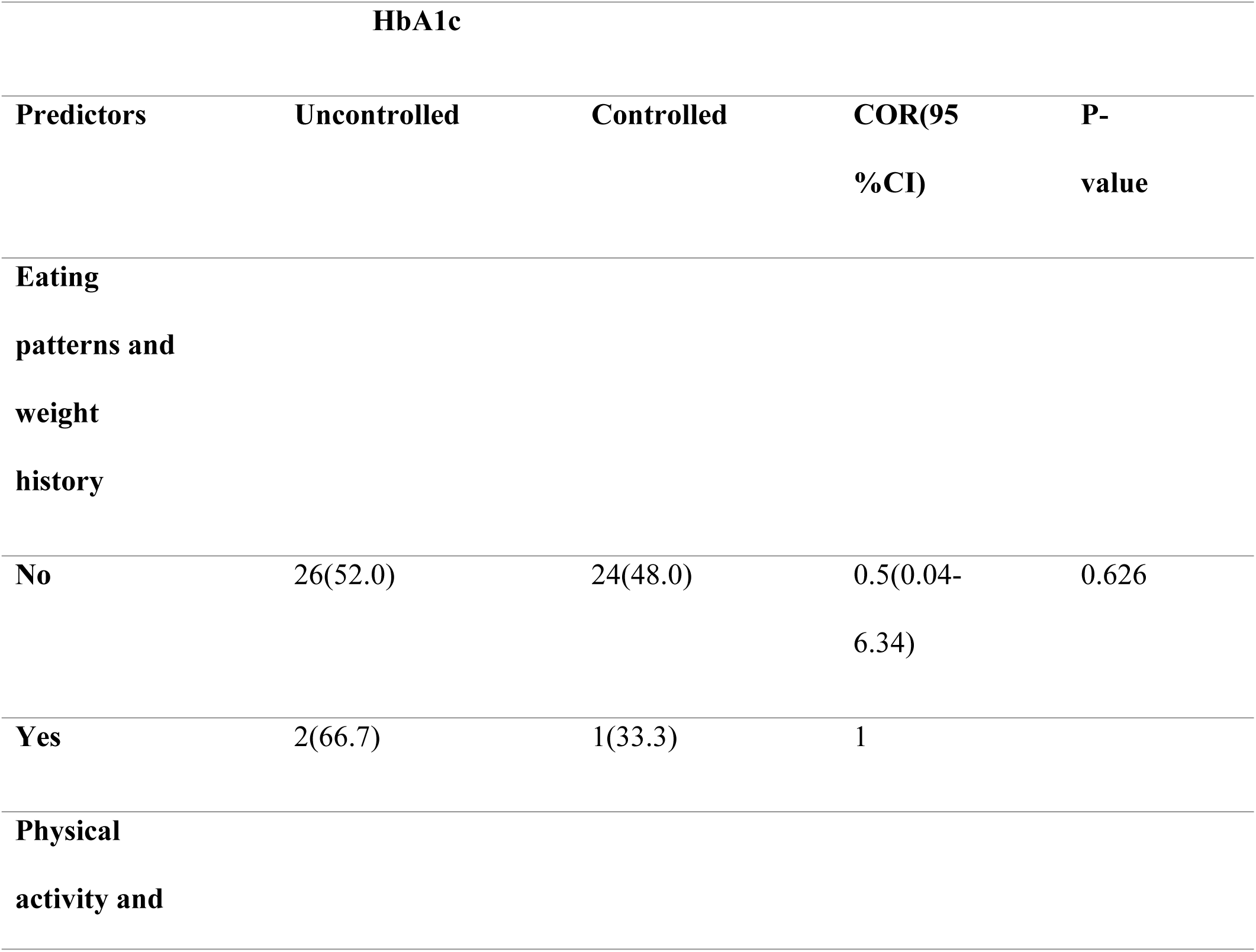
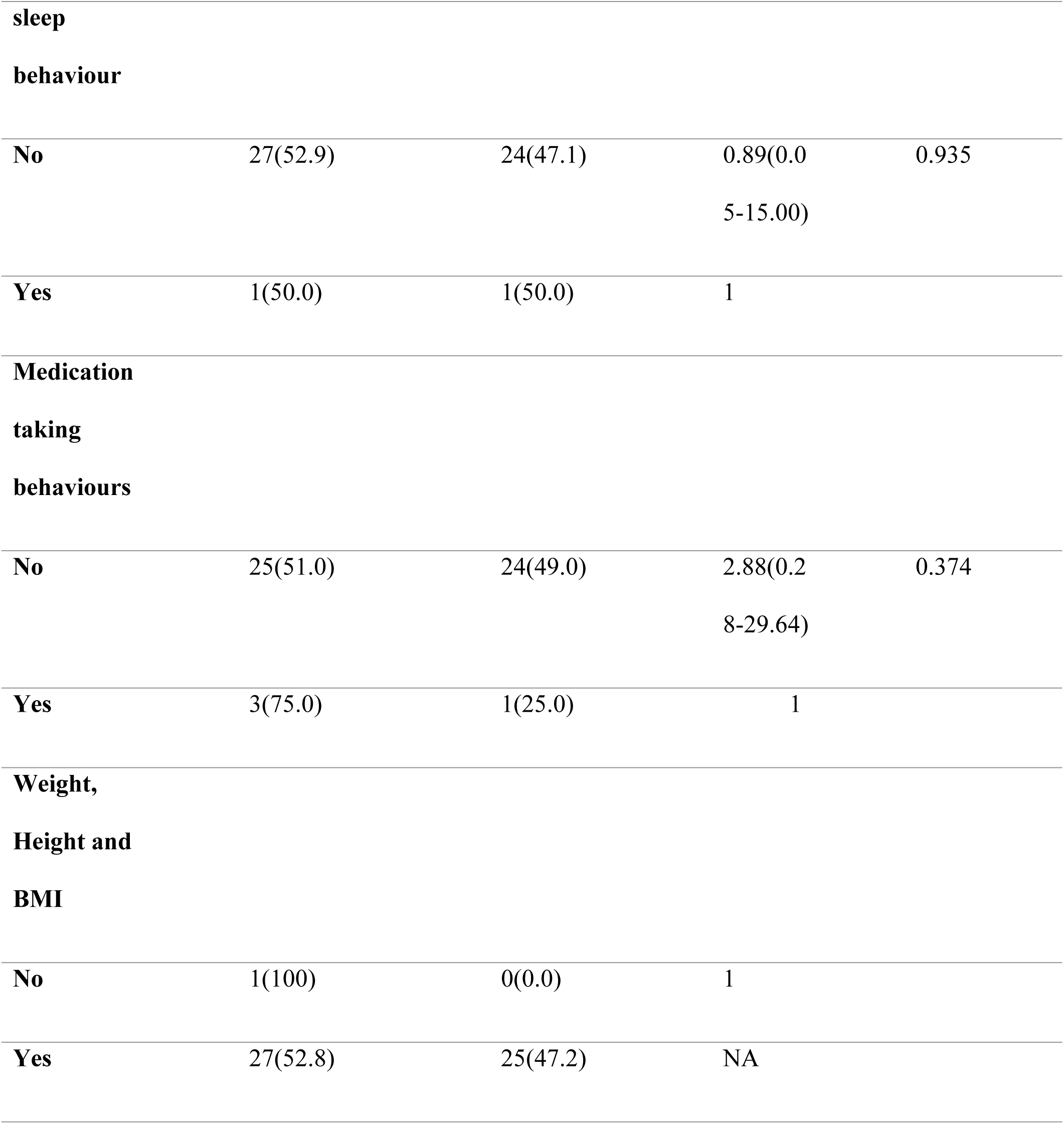
Predictors of HbA1c

## Discussion

The overall documented adherence rates in this study were 17%, 58.4%, 24% and 5% for medical history, physical examination, laboratory evaluation, and referrals, respectively. These were coupled with low rates of achievement of treatment targets for various clinical outcomes, specifically glycated hemoglobin (HbA1c), blood pressure, lipid profile and estimated glomerular filtration rate (eGFR), with corresponding percentages of 47.2%, 42%, 47.1%, and 48.8%, respectively. Looking at the overall mean documented adherence to ADA standards, the current study shows that approximately 72% of patients were not managed according to the ADA standards of care, strictly considering the documentation done on the electronic medical record system, with reference to guidelines for follow-up visits and annual visits.

The older age group of 60 to 80 years had the highest frequency amongst the records viewed, and a similar pattern was shown in a community-based prevalence study done in Accra, where the highest diabetes prevalence rate was recorded amongst the oldest age group (64+ years). DM is a very common phenomenon amongst older persons with various age-related changes contributing to insulin resistance. Aside from its microvascular and macrovascular complications in the elderly, it also further contributes to decreased quality of life, cognition, tolerance of pain, and increased risk of falls, nocturia, pressure ulcers, amongst others, which necessitates that good glycaemic control be achieved in this population by comprehensive evaluation and care.

The overall documented compliance rate for history taking was 17% which is low. The compliance rates for all parameters under history taking were lower than 10% with the exception of the current medication regimen, which suggests very poor compliance in that regard. There are no similar studies within the country to compare these findings to, but a similar study done in Saudi Arabia from various primary care clinics showed a higher percentage of compliance (44.86%), even though the elements under history taking were more than what was considered in this study. Eating habits and physical activity patterns had compliance rates of approximately 59% which is way higher than what has been found in a tertiary care facility in Ghana [14]. The sub-elements under history taking per the ADA guideline help to evaluate each individual patient’s condition thoroughly to be able to assess and advice on the needed lifestyle modification as well as medication adherence. The fact that the documented adherence to such a crucial aspect of patient care is low calls for urgent efforts to bring about the needed improvement.

The overall documented compliance rate for physical examination was relatively higher at 58.4% which is the highest percentage recorded amongst all the categories assessed. Findings from this study are similar to those recorded in a similar study in Saudi Arabia, where the compliance rate for weight, height, and BMI was about 94% and 100% for blood pressure measurement, and that of skin examination was 4.5% which is higher than what this study recorded [14]. Obesity is a significant risk factor for the development of diabetes and research has shown that weight loss of 5% or more of total body weight is associated with improvements in glycaemic control, the decreased need for diabetes medications, and improved quality of life and so weight monitoring and BMI calculation is important in the management of diabetic patients (15). Blood pressure monitoring is also important during patient care as people with diabetes are at risk of elevated blood pressures which coupled with the hyperglycaemic state in DM increases the risk of cardiovascular complications and consequently worsens the rate of mortality [16]. There was a wide disparity between the high documented adherence rate recorded in blood pressure measurement (96%) and the relatively low clinical outcomes of well controlled blood pressures (41.9%) and the logistic regression analysis did not reveal blood pressure measurement as a significant predictor of blood pressure control (P=0.599) which suggests that beyond blood pressure monitoring, adequate control depends on other factors as well. A similar study done in Korea showed a similar trend where the adherence rate for blood pressure measurement was about 94% while achievement of target blood pressure was about 28% [17]. A cross-sectional multicentre study carried out in a developing country also showed a similar finding [18].

Overall documented compliance rate for referrals was the lowest (5%). Lifestyle modification including increased physical activity and diet control is very important in controlling risk factors such as obesity in management of DM and that is why patients ought to be referred to a dietician, but the performance rate is so poor from our study, and this may contribute to poor glycaemic control. Eye screening is also key in diabetes management because diabetic retinopathy is one of the microvascular complications that can eventually lead to loss of vision and hardly shows any symptoms in the early stages until it becomes advanced, making screening important for early detection of retinopathy, which can be managed easily than in advanced stages [19]. Unfortunately, the documented compliance rate for eye screening too was not good.

According to a qualitative study done in Ghana on the post-implementation challenges of the electronic health record system, negative attitudes of staff towards the judicious use of the record system were a major one, possibly due to frustration with poor network and connectivity [20]. This is to say that the electronic health records system has its own challenges which can affect the accuracy of objective research work that depends solely on documentation as seen in this study.

There was no documentation of any referral made to a certified diabetes educator for diabetes self-management education and support, and that is because there are currently two certified diabetes educators, and they are the same nursing staff who perform the routine clinic services on a daily basis. Diabetes self-management education and support is carried out as part of their routine services at the clinic. The compliance rate for the element of annual eye exams in a study in Saudi Arabia for primary care clinics was higher (63%) than what this study recorded. The overall compliance rate for referrals in the same study, even though not a good performance (19.33%), was relatively better than that in this study as well [14]. Adherence rates for referral for annual eye examinations in various studies done in Nigeria and Egypt were relatively higher than what this study recorded with 48% and 64.5% respectively [21,22].

Laboratory evaluation had the second-highest overall compliance rate (24%), which is not a good performance. The performance of various primary care clinics in a study done in Saudi Arabia was relatively higher (36.28%) but still not up to even 50% [14]. However, the differences in the level of health facilities in comparison here makes the case in Ghana a more serious one.

Glycated haemoglobin (HbA1c) measurement is used to assess glycaemic control and is commonly used in clinical trials to demonstrate the benefits of improved glycaemic control. The ADA recommendation is that it should be assessed at least 2 times a year in patients who are meeting treatment goals, and for this study, we looked out for whether it was done at least once a year and even with the lower standard set, the documented compliance rate for HbA1c measurement was 42.6% which is not even up to 50%. In a similar study done amongst type 2 diabetic patients in primary care at Saudi Arabia, the adherence to ADA process standards of measurements of HbA1c was 68.7% [23].

The clinical outcome of a well-controlled glycaemic status was about 47% for those whose results were readily available which means that good glycaemic control was not achieved for more than half of the patients and the low documented compliance rate could account for this to a great extent. However, in the research done on type 2 diabetes patients, it was noticed that even though a relatively higher adherence rate of 68.7% was recorded, only 24.2% of patients had a well-controlled glycaemic status which means that the rate of achievement of target is lower than the adherence rate. The contrast in findings could be due to the fact that for this study, the percentage of patients whose results were readily available and used for assessment of clinical outcomes was very low (20.7%). Regression analysis did not reveal HbA1c measurement as a significant predictor of good glycaemic control from this study (P=0.69). Moreover, several shortcomings with the operation of the electronic medical record (EMR) system which negatively affected documentation, suggests that the study results may not fully justify the strength of the conclusions.

In a similar study done in Nigeria, only 14% of patients had their HbA1c measurement done within the year of review with a mean HbA1c clinical outcome of 10.5+2.0%[22] while the mean HbA1c value from this study was 7.72%. It is very evident then that their poorer adherence rate resulted in them recording a higher average mean HbA1c than ours which signifies uncontrolled hyperglycaemia in that population relative to ours. A similar trend was noticed in the mean fasting plasma glucose level recorded in this study and theirs. That recorded in this study was 8.10 ± 3.40 mmol/l while that recorded in their study was 9 + 1.13 mmol/l.

Dyslipidaemia is simply the imbalance of lipids such as triglycerides, LDL-cholesterol, HDL-cholesterol amongst others and diabetic dyslipidaemia is characterized by elevated fasting postprandial triglycerides, low HDL-cholesterol, elevated LDL-cholesterol and predominance of small dense LDL particles and this tend to demonstrate the link between diabetes and the increased cardiovascular risk in DM patients [24]. The annual lipid profile measurement is therefore a very important standard process recommended by ADA. The documented adherence rate this study was 49.1%, while an adherence rate of 80.2% was recorded in a similar study for type 2 diabetics [23]. A similar study in Lagos also revealed a higher adherence rate of 61% [22]. Considering the serum lipid panel as an outcome measure, approximately 53% had an abnormal serum lipid profile. This calculation should be viewed from the background of the availability of only 26.5% of results out of the entire number of records viewed. Therefore, only 47% of the patients whose results were available achieved serum lipid targets.

Diabetic kidney disease is a clinical diagnosis clinicians make based on the presence of albuminuria and or decreased eGFR in the absence of signs and symptoms of other primary causes of kidney damage. ADA therefore recommend an annual serum creatinine/eGFR determination. The adherence rate was approximately 53% and for the clinical outcomes, only about 48.8% of patients whose results were available had normal eGFR values (G1-G2) and this suggests that the prevalence of renal complications is high and will get worse if standards are not adhered to. Adherence to screening was relatively lower (35.6%) in the study done in Saudi Arabia as compared to this study [23].

The compliance rates for urinary albumin to creatinine ratio, thyroid function test, vitamin B12 and serum potassium were all less than 15%. According to the electronic records, nobody on metformin was made to do a vitamin B12 test and thyroid function test had a compliance rate of only 2%. This could be due to the fact that the test is primarily performed for those with type 1 DM whose population is relatively smaller as compared to the type 2 DM population.

Findings from this study clearly shows that the documented adherence rates generally to ADA process standards on history taking, laboratory investigation, physical examination and referrals at CCTH are suboptimal and that there are quite a number of gaps in the care of diabetic patients. It is also evident that all the clinical outcome targets were not achieved in majority of patients which is a reflection of our poor management standards. However, other studies that had better adherence rates to some of these ADA standards as has been clearly shown in previous paragraphs did not really see a perfect correlation in terms of better outcomes. Another clear example is in a study done in Egypt which showed an adherence rate of about 61% to HbA1c measurement in the previous 3 months within the period of study but revealed uncontrolled hyperglycaemic states in about 99% of cases [25]. This is not the same for all similar studies done though. Some studies have also shown with statistical significance that good adherence rates lead to better patient outcomes and reduces mortality. For instance, a study done in Korea revealed that patients that lacked data or information on their blood pressure, HbA1c, eye examination and LDL cholesterol amongst others had worse outcomes which specifically were end stage renal disease and death. It also revealed that there is a statistically significant relationship between receiving eye examination, optimal glycaemic control and better clinical outcomes [17].

These findings together reveal the complex nature of diabetes mellitus and its multifaceted components which suggests that good adherence to certain standard processes in clinical care can only partly contribute to better clinical outcomes. Research has shown that improvement in clinical outcomes such as HbA1c do not always follow improvement in the processes of care as patient factors such as adherence to dietary advice and moderate exercises, comorbidities, safety and cost of medications all work together to influence patient outcomes [25]. Socioeconomic factors such as financial constraints really influence management outcomes as it is one of the major reasons stated in the records viewed as to why patients were not doing the routine labs ordered for them in this study considering our setting where these tests have to be paid for. Patient default from clinic attendance was also another issue picked up in the records from this study that tends to contribute to poor glycaemic control and clinical outcomes. Nonetheless, good adherence to clinical guidelines is considered a major step towards the achievement of better patient outcomes. There is therefore the urgent need for clinicians and all health workers to be diligent and responsible in adhering to standards for comprehensive evaluation and management of patients and to document appropriately to help make evaluation of systems easier and more accurate.

Type 2 diabetes is associated with a greater incidence of liver function test abnormalities relative to those who do not have DM and chronic mild elevation of transaminases are frequently found in this category of patients which reflect some underlying insulin resistance [26] as well as non-alcoholic fatty liver disease in some patients with type 2 diabetes mellitus [27]. Adherence rate from this study to annual liver function test was about 24% and only 36% of patients achieved the expected target results for the various specific markers in the liver function test panel. It should be noted that the percentage of records viewed who had available liver function tests to be assessed was only 14% indicating the need for cautious interpretation of results. These percentages and markers suggest that there is a gap in the standard of care that must be filled and made better.

### Limitations of the study

This study had its own limitations. The quantitative study design employed did not really consider the context and the local conditions in which the study was done but efforts were made to engage some staff for their input which has been shared in the paragraphs above. Then also, the generalizability of the study is limited as the study population was strictly patients from the teaching hospital and so the findings cannot be applied to the general diabetes population.

Our analysis was based on the ADA 2022 guidelines in effect at the time, which endorsed a blood pressure target of <140/90 mmHg for the average adult. Subsequent revisions in the ADA 2023 guidelines lowered the recommended target to <130/80 mmHg, differing from the threshold originally applied.

Then also, the fact that the hospital had issues with the electronic medical record (EMR) system prior to it being fixed in 2023 suggests that clinicians may have had difficulty with documentation. Consequently, the conclusions from the study might be overstated. Moreover, there could have been some selection and documentation biases which can affect the validity of the findings of the study.

## Conclusion and recommendations

In conclusion, the management processes in the diabetic clinic showed low rates of documented adherence to ADA standards for medical history, physical examination and laboratory evaluation but physical examination was relatively better than the others. This study hypothesized that there was a high documented adherence to ADA standards with no significant difference or gaps, but the findings proved otherwise with gaps seen in history taking. Skin examination was a significant gap in physical examination and for laboratory evaluation there were gaps in the ordering of HbA1c, lipid profile, renal function tests amongst others. Patient’s results were not being uploaded on the system. There were great gaps in referrals for eye examination and to dieticians as well. However, these findings should be interpreted with caution due to the challenges with the hospital’s EMR system. The evidence may be insufficient to support the extent of the conclusions.

Diabetes is a dangerous epidemic on the rise that demands utmost attention and findings from this study have revealed the gaps and setbacks. These recommendations below will help address these gaps in our quest to combat this disease as a nation.

Clinicians and all health workers must be diligent and responsible in adhering to standards for comprehensive evaluation and management of patients and document appropriately on the electronic medical record (EMR) systems to help make evaluation of systems easier and more accurate. Training on appropriate use of the EMR should be organized for staff at health facilities and the importance of complete documentation should be emphasized during such training.

Diabetes education is key and such education campaigns should be extended to the public beyond what is done by doctors and diabetic educators in the walls of the hospital to help reduce patient default. More diabetic nurses and diabetic educators should be trained to help in diabetes self-management education and support.

The Ghana government and the Ministry of Health must build a robust system where logistics are readily available and work at making routine tests for patients highly subsidized to help patronage.

## Data Availability

The data supporting the findings of the study shall be held in a public repository specifically, figshare, and this information will only be available after acceptance of manuscript to all appropriate and relevant stakeholders without restrictions.

https://figshare.com/s/60f1bbfa4420422ca44b

## Acknowledgments

I also want to acknowledge and appreciate the efforts of Dr. Kanyike Andrew Marvin, Drs. Lily Owusu Frimpong and Kofi Andoh, whose contributions and guidance have been so crucial to this research.

